# Upright Participation for Learners with Physical Disabilities: A Pilot Randomized Crossover Effectiveness-Implementation Trial of Robot-assisted Walking in Schools

**DOI:** 10.64898/2026.10.16.26365549

**Authors:** A Hilderley, C Diot, B Morgan, E Byrne, E Chu, A Gosselin, K Sheldon, A McCormick, FV Wright, L Pritchard, H Shen, A Kirton, EG Condliffe

## Abstract

**Background:** School-based implementation of robot-assisted walking may support increased physical and social participation for children with physical disabilities, but evidence from an adequately powered trial is needed. This pilot study evaluated implementation strategies and study procedures to inform a future definitive trial.

**Methods:** Participants were randomized to 12-weeks of robot-assisted walking or control, with subsequent crossover to the other condition. In the intervention arm, targeted dose was 75 minutes of robot-assisted walking, spread across three days/week. Implementation strategies for the child’s classroom team included educational materials, centralized technical assistance, school champions, and on-going training. Implementation strategy evaluation focused on intervention protocol feasibility (i.e., weekly device use, troubleshooting) and fidelity (documentation of implementation strategy modifications). Preliminary estimates participation goal changes were obtained using the Canadian Occupational Performance Measure (COPM) ‘Performance’ scale (/10).

**Results:** Eight children with neurodevelopmental physical disabilities participated (mean age 3.5, SD 0.5 years). Device use varied from 2.1 to 4.3 days/week (median 3.3) with a range of 48 to 96 minutes/week (median 80). Target use of ≥ 3 days/week was achieved for 66% of weeks and met the ≥75-minute target for 33% of weeks. Troubleshooting occurred in 18.5% of sessions. Two implementation strategies were modified to improve feasibility. COPM mean change was 3.56 (SD 1.07) for robot-assisted walking and 2.11 (SD 2.41) for control (d_z_=0.74).

**Conclusions:** This pilot study provided preliminary evidence regarding both the feasibility of robot-assisted walking implementation in schools and its potential effectiveness for children with physical disabilities. Findings support progression to a definitive trial.

## Introduction

More than one in every 1000 children have a neurodevelopmental disability that leads to significant mobility limitations and impacts their ability to participate in meaningful life activities.^1–4^ Upright participation has the potential to influence multiple aspects of these children’s development and daily life.^5^ Despite the large need, there are few, if any, interventions available to facilitate upright participation and motor function for children with significant mobility limitations who are primarily wheelchair users.^6^

Recently, robot-assisted walking has been shown to improve motor function and reduce the health impacts of physical inactivity that these children often experience.^7–10^ Robot-assisted walking devices attach to a walker and use robotic legs to power a child’s leg movements, allowing them to use their body in new ways in an upright weightbearing position. When used in home and community settings, it also creates new avenues for participation in daily life that are critical for supporting cognitive, communication and social development.^9^ However, this technology is expensive and requires adult support, limiting access to families with adequate resources. Equitable opportunities to access robot-assisted walking are required to improve multidimensional outcomes for non-ambulatory children.

Previous studies have identified school settings as an optimal context for equitable access to robot-assisted walking to promote participation and function-based improvements.^9^ To facilitate successful implementation in schools, it is important to systematically identify barriers, facilitators, and strategies for implementation.^11^ As a first step, we conducted a pilot study to identify implementation strategies for robot-assisted walking in school settings.^12^ Barriers and facilitators were mapped to constructs of the Consolidated Framework for Implementation Research (CFIR)^13^ and matched to implementation strategies from the Expert Recommendations for Implementing Change (ERIC) compilation.^14^ Researchers and school physiotherapists then collaboratively reviewed the matched strategies and selected 14 that best addressed identified barriers and leveraged facilitators.^12^

Before conducting a fully powered, multisite randomized control trial, implementation strategies should be piloted within the planned hybrid type 2 effectiveness-implementation trial design.^15,16^ Piloting hybrid type 2 designs supports evaluation of potential implementation strategy effects in addition to assessment of whether the protocol methods are feasible to replicate in a definitive trial.

Our school-based pilot study had two primary objectives. The primary implementation objective was to assess potential implementation strategy effects, with a focus on feasibility and fidelity of implementing robot-assisted walking in schools.^15^ The primary effectiveness objective was to obtain preliminary estimates of change in individualized, school-based participation goals to inform the design and sample size of a future definitive trial.

## Methods

This pilot effectiveness-implementation study^15,16^ employed a prospective, open label, randomized (1:1) crossover controlled (AB/BA) design, thus providing equitable intervention access to all participants.^17,18^ The trial had pragmatic intentions.^19^ The ethics committee of the University of Calgary (Calgary, Canada) approved the protocol (REB24-0305), and the study was registered on clinicaltrials.gov (NCT07726706).

Reporting follows relevant reporting guidelines, specifically StaRI for implementation studies,^20^ the CONSORT extension for pilot and feasibility trials,^21^ and GRIPP2-SF for reporting of patient and public engagement on our research team.^22^ Our research team included two family partners (AG, KS) and two physiotherapist knowledge users (EB, EC). We collaborated on trial design, with their input being particularly beneficial for increasing the real-world applicability of the study.

### Participants & Settings

Participants attended special education schools: Pacekids (Calgary, Canada) or Renfrew Educational Services (Calgary, Canada) which had two locations. We intentionally included three locations to learn whether implementation strategies could be successfully tailored to different contexts.

The target sample size was eight participants, chosen to support the randomization scheme.

Eligible children were aged two to 10 years old with neurodevelopmental disabilities who were primarily wheelchair users and could reliably indicate discomfort to school staff. Maximum child size was determined by whether school staff could safely support transfers into the device. Diagnostic criteria were not specified, as previous research found no differences in outcomes between children with cerebral palsy and other diagnoses, including progressive conditions.^7^ Exclusion criteria were: (i) medical conditions or recent surgery requiring lower-extremity immobilization or weight-bearing restrictions; (ii) medical conditions requiring physical activity restrictions; (iii) pain or symptomatic hypotension while standing; or (iv) contracture preventing forward movement in the device.

Recruitment was organized through the two partner schools. Before the end of September 2025, each school identified children eligible for the study and obtained consent from families for researcher contact. Research team members then screened participants over the phone. If eligible and interested, the child had a test session in the robot-assisted walking device before enrolment to ensure fit and comfort. Informed consent/assent was electronically obtained from guardians.

### Randomization

Participants were randomized 1:1 within each school to begin with robot-assisted walking or waitlist control, followed by crossover to the alternate period (AB/BA design). Randomization used permuted blocks of two, with children requiring the same device size grouped within blocks to ensure device availability. Allocation was concealed using a centralized, computer-generated randomization sequence generated by a blinded investigator (EGC). The allocation sequence was inaccessible to study personnel responsible for participant enrollment until pre-Period 1 assessments were completed. The trial was open label to support monitoring of study methods.

### Treatment

The intervention and control periods were each 12 weeks. During the control period, participants used their usual mobility devices at school (e.g., wheelchair, walker, standing frame). During the intervention period, participants used the Trexo robotic walker (Trexo Robotic, Mississauga Canada) as their mode of mobility and/or as a standing aid. The Trexo attaches to a walker, which provides a stable base of support. Motorized joints at the hips and knees guide a child’s legs through a stepping pattern overground, with adjustable levels of robotic assistance to accommodate each child’s abilities. There are four device sizes, with selection based on a child’s anatomical measurements. During the brief set up time of approximately 5-10 minutes, a device operator transfers the child into the device and secures its adjustable straps to their legs. The device operator controls the device using a dedicated tablet attached to the device.

School physiotherapists and support staff with dedicated 1:1 time with participants received training to be the device operators. They completed the virtual training process from Trexo Robotics, which has been successfully used in previous trials.^8,10,12^ Training included individualized adjustment of the device to each participant, instruction in safely operating the tablet and selecting appropriate gait settings, and virtually supervised practice using the device with their school participant.

Trexo device use occurrences and activities were not completed according to a prescribed program. Instead, school staff were encouraged to use the device with the participating child as much as possible during the school day, with a target of 3 days and 75 minutes of accumulated use per week. Usage data, specifically number of times used, duration of use (minutes), and number of steps, were documented automatically on the tablet connected to the device. Device operators also completed a simple user log in Microsoft Excel each week to detail barriers, facilitators, and reasoning for weeks that had lower or higher than targeted usage.

### Implementation Framework and Strategies

The Quality Implementation Framework (QIF) guided the implementation process across four phases.^23^ For the first phase, “initial setting considerations”, three strategies were selected from our previously identified list of implementation strategies and enacted as follows:^12^

- *Involve recipients and family members*: Our implementation team included parents of a Trexo user (AG, KS).
- *Conduct local needs assessment*: Local needs assessments were conducted through informal meetings between the research and school teams. Discussions considered the number of potentially eligible participants, required device sizes, available school staff to be device operators, and how use of devices could fit within the school day.
- *Assess for readiness and identify barriers and facilitators*: Planning meetings were conducted with the school teams to determine the unique needs specific to each site. Study timelines and processes were reviewed. School teams shared anticipated facilitators and potential barriers, which informed selection of subsequent strategies.

During the second QIF phase, “creating an implementation structure”, school teams selected strategies from our list.^12^ Strategies were tailored to their site as follows:

- *Develop educational materials*: A troubleshooting guide was created and shared with school teams. The extensive experience of the research team was used to inform both the issues and solutions detailed in the guide.
- *Centralized technical assistance*: School teams were connected with the Trexo Robotics company, which has a messaging application for troubleshooting issues related to software, connectivity, or participant setup.
- *Identify and prepare school champions*: School physiotherapists (n=4) and a therapy assistant (n=1) were identified as site champions, with at least one per school site. Three of the four physiotherapists had previous experience with the Trexo. Champions served as the primary point of contact for any study or device use questions.
- *Promote adaptability*: Conversations with school leadership ensured that school staff were offered the freedom to organize responsibilities amongst themselves to provide the necessary flexibility to decide when and who operated the device within their busy school context.
- *Conduct on-going training*: Throughout the study period, the research team and school staff remained in constant communication to efficiently address barriers in real-time. A dedicated research team member was available for advice on participant setup or replacing hardware.

The third QIF phase, “ongoing structure once implementation begins”, involved monitoring and adapting strategies as needed (see Outcomes section for details). Finally, phase four of “improving future applications” focused on informing the future definitive trial. We detail successes and challenges in the Results and Discussion sections.

### Implementation Outcomes

We set benchmark criteria for implementation outcomes, specifically Reach, Adoption, Feasibility, Fidelity, Safety, and Sustainability.^24,25^ These criteria assessed methodology to be retained or revised in our future definitive trial. Under Reach, we assessed recruitment and attrition. Adoption considered the number of device operators per participant and the roles of trained device operators (e.g., educator, aide, physiotherapist). Feasibility considered weekly device use, troubleshooting instances, and assessments. Fidelity had a particular focus on the Framework for Reporting Adaptations and Modifications to Evidence-based Implementation Strategies (FRAME-IS) to document adaptations and modifications to implementation strategies,^26^ along with completion of device usage logs. Adverse events were tracked for safety. Finally, for sustainability, we held an end-of-study meeting between research and school teams to identify methods to retain or change in the future definitive trial.

### Effectiveness Outcomes

Assessments were conducted at three timepoints: pre-period 1 (pre-randomization), post-period 1/pre-period 2, and post period 2 (see Figure 1). These were completed by a school physiotherapist at the participant’s school or by a research clinician from the Alberta Children’s Hospital (Calgary, Canada).

**Figure 1.**
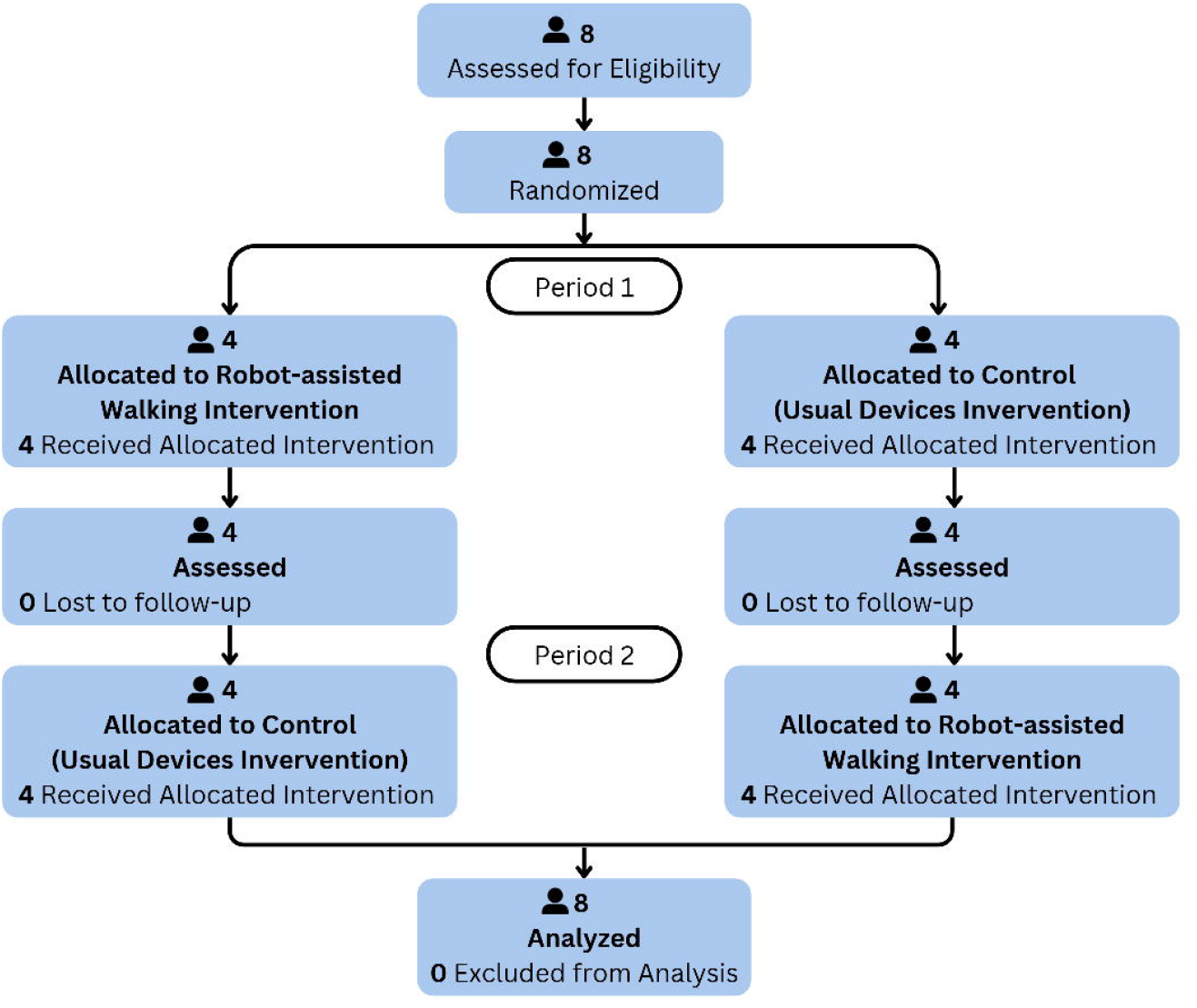
Participant flow through the trial. **Caption**: Period 1 was completed from October 2025 to January 2026, with the school winter holiday between Week 9 and 10. Period 2 was completed from February 2026 to June 2026, with the school spring holiday after Week 2, 4, or 5 (participant and site dependent).

The primary effectiveness outcome measure was the Performance scale of the Canadian Occupational Performance Measure (COPM).^27^ Before period 1, participants collaboratively identified 3–5 goals related to social or physical participation or motor function with their school physiotherapist, which were reviewed with families by phone or virtually. A trained research clinician (CD) then guided the device operator most involved with each participant (school physiotherapist or aide) to rate goal Performance and Satisfaction on a 1 to 10 scale. The same respondent rated goals at post-period 1/pre-period 2 and post period 2 using the same procedures. As well, each COPM goal was reviewed after period 1 by each participant’s physiotherapist or aide who worked with them most closely. If there appeared to be potential for a scoring ceiling effect based on their judgement, descriptors were updated and rescored before period 2.^28^ Performance and Satisfaction (secondary outcome) mean scores (across goals) were calculated at each timepoint. A change of ≥2 points is commonly considered clinically important, although the threshold is context dependent.^29^

Goal Attainment Scaling (GAS) objectively quantified goal achievement on a 5-point scale from -2 (baseline score) to + 2 (much better than expected).^30,31^ GAS goals were selected and scored by a trained research clinician (CD) by week 2 of each period. For each GAS goal, an outcome measure was selected that best matched that goal. To assess whether the definitive trial could be decentralized, i.e., all measures could be completed at the children’s school rather than a central site, preference was given to validated measures that could be completed online or videotaped by school staff for subsequent scoring.

Selected validated measures that could serve as a foundation for setting GAS outcome achievement levels included the Young Children’s Participation & Environment Measure (YC-PEM)^32^ to assess goals related to participation frequency, the Early Clinical Assessment of Balance (ECAB)^33^ to assess static and dynamic postural control goals, and the two-minute walk test (2MWT)^34,35^ to assess walking endurance goals. When these measures did not align with the COPM goal, individualized tasks were chosen for the GAS goal. GAS scaling was based on the scoring characteristics of each measure and clinical judgment of the school physiotherapist. See Supplemental Material for additional details. Post-intervention GAS raw scores (-2 to +2) were converted to an overall T-score for analysis, with a T-score ≥50 indicating (as is standard practice for GAS use) that post-intervention change met or exceeded expectations.

### Statistical Analysis

Implementation outcomes were summarized using descriptive statistics appropriate to the measure and compared with pre-specified benchmark criteria. Device use was calculated across all intended weeks, excluding weeks with planned non-use (e.g., school holidays, student absence). Depending on variable type and distribution, mean, SD, median, IQR, and range were reported. Effectiveness outcomes were summarized descriptively as mean change (SD) within each condition and as the within-participant difference between conditions with 95% confidence intervals. Consistent with guidance for pilot trials, no hypothesis tests of effectiveness were performed.

COPM Performance scores were used to inform sample size calculations for the future definitive trial. Treatment contrast, paired Cohen’s d, and between-condition correlation were calculated, and period and carryover effects were examined descriptively by comparing, between the two sequence groups, the within-participant differences and the within-participant sums of the two period change scores, respectively. Given four participants per sequence, these comparisons were not used for inference. Sample size was estimated using a paired t-test reflecting the AB/BA crossover design, with two-sided α=0.05 and 90% power. Sensitivity analyses varied the assumed effect size and between-condition correlation. A Monte Carlo simulation with 1000 trials was then used to verify adequate power under a planned multisite mixed-effects model and to assess the impact of between-site heterogeneity in treatment effects. Simulations reflected the planned AB/BA crossover design, with children allocated approximately equally across 10 sites and randomized 1:1 to sequence.

## Results

Eight participants (two female) were enrolled in the trial, with a mean age of 3.5 (SD 0.5) years. Figure 1 depicts trial flow. The first participant was enrolled on October 6, 2025, and the final assessment was completed on June 16, 2026. Most participants had a diagnosis of cerebral palsy (75%), and all were primarily wheelchair users. Participant characteristics are detailed in Table 1.

**Table 1.** Participant Characteristics.

| Characteristic | Sequence: Active-Control | Sequence: Control-Active | Total Sample |
| --- | --- | --- | --- |
| Sample size (n) | 4 | 4 | 8 |
| Age (mean, SD) | 3.54 (0.60) | 3.46 (0.48) | 3.50 (0.50) |
| Sex (n) | 3 male<br>1 female | 3 male<br>1 female | 6 male<br>2 female |
| Primary Diagnosis (n) | 3 cerebral palsy<br>1 genetic condition | 3 cerebral palsy<br>1 genetic condition | 6 cerebral palsy<br>2 genetic condition |
| GMFCS Level (n) | 2 Level IV<br>1 Level V<br>1 N/A | 3 Level IV<br>0 Level V<br>1 N/A | 5 Level IV<br>1 Level V<br>2 N/A |
| School site (n) | 2 Site A<br>1 Site B<br>1 Site C | 1 Site A<br>1 Site B<br>2 Site C | 3 Site A<br>2 Site B<br>3 Site C |
| Device size (n) | 2 Medium/Large hybrid<br>1 Medium<br>1 Small | 2 Medium/Large hybrid<br>1 Medium<br>1 Small | 4 Medium/Large hybrid<br>2 Medium<br>2 Small |

**Legend:** Active: robot-assisted walking intervention; GMFCS: Gross Motor Function Classification System; N/A: not applicable to children with a genetic condition; School sites A and B: within the same school system of Renfrew Educational Services; School site C: Pacekids.

### Implementation Outcomes

As detailed in Table 2, 11 of 13 implementation indicators met criteria.

**Table 2.** Implementation Outcomes.

| Outcome Area | Indicator | Criteria | Results |
| --- | --- | --- | --- |
| Reach | % identified who participated<br>% attrition | 90%<br><10% per period | **100%**<br>**0%** |
| Adoption | # trained staff<br>Roles of trained staff | ≥2 per site<br>≥2 designations | **≥4 per site**<br>**6 designations** |
| Feasibility | Weekly device use ≥75 mins<br>Weekly device use ≥3 days<br>Troubleshooting instances<br>Complete assessment data | 80%<br>80%<br>≤30% of sessions<br>90% | **33%**<br>**66%**<br>**18.5%**<br>**100%** |
| Fidelity | Documentation using FRAME-IS<sup>26</sup><br>Log completion | 100% complete<br>>80% complete | **100%**<br>**100%** |
| Safety | Minor or moderate adverse events<br>Severe adverse event | <10% sessions<br>0 total | **0.74%**<br>**0** |
| Sustainability | End-of-study review of methods | 1 meeting | **Completed** |

**Legend:** FRAME-IS: Framework for Reporting Adaptations and Modifications to Evidence-based Implementation Strategies; **green text**: criteria met; **red text**: criteria not met.

#### Reach

All eligible children were enrolled and completed the trial. Randomization methods were successful with no attrition in either period (see Figure 1).

#### Adoption

Multiple school staff were trained as device operators at each site, with designations including school physiotherapist, therapy assistant, physiotherapy student, program coordinator, aide, and preschool teacher.

#### Feasibility

All eight participants completed the 12-week robot-assisted device intervention. Four participants had 1-3 full weeks (8 weeks in total) of unavoidable non-use due to school absence or temporary activity restrictions (see Fidelity below for details). With these weeks of non-use omitted, median (range) usage was 3.3 (2.1 to 4.3) days/week, 79.7 (48.4 to 96.1) minutes/week and 3,202 (1,867 to 3,947) steps/week.

Across participants, there were 58 weeks with ≥3 days of use (66%) and 29 weeks with device usage ≥75 minutes (33%). All participants used devices for ≥3 days for at least three weeks (median 7.5 weeks, range 3 to 10). Seven of the eight participants used devices for ≥75 minutes for at least one week (median 3.5 weeks, range 0 to 8). Figure 2 depicts device use and shows that device use did not decline over time.

**Figure 2.**
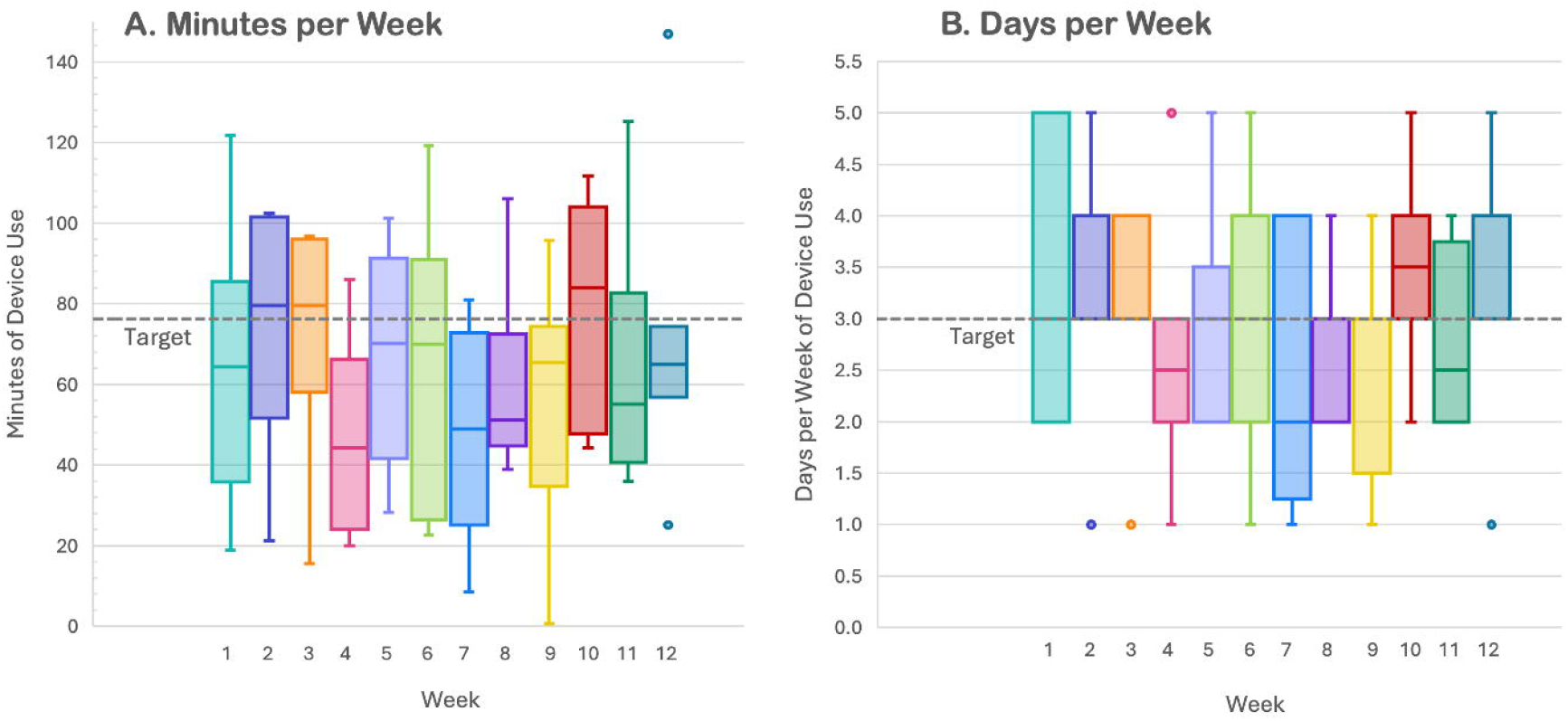
Robot-assisted Walking Device Use. **Caption**: Device use per week for (A) Minutes (B) Days. Boxes represent the interquartile range (25^th^-75th percentiles), centre lines indicate medians, and whiskers extend to the most extreme values within 1.5 times the interquartile range. Points beyond the whiskers indicate outliers.

Of the 270 total sessions, there were 50 reports of device problems (18.5% of sessions). More than half of device problems were Wi-Fi connection issues (10.4%) Either it took longer than expected to connect the tablet to the device or connection was lost during a session which required reconnection. Other device problems included batteries requiring charging, setup issues and adjustments (e.g. adjusting gait settings for improved toe clearance, adjusting straps to better secure feet, adding hip belt).

All assessments were completed as planned, confirming that in-person assessment components could have been completed in each child’s school environment.

#### Fidelity

Logs were returned for all participants, with comments for each weekday or for each day with intended use. Logs reported that a total of 70 planned device use sessions were missed. Participant absence from school or medically advised activity restrictions (e.g., following a medical procedure) accounted for 58 (82.9%) of these missed sessions. For the remaining 17.1% of missed sessions, 5 (7.1%) reported missed sessions due to school logistics, including lack of time or the main device operator was sick and there were no other staff available. There were 4 (5.7%) sessions missed as a result of a connection issue (unable to login on the tablet or no internet at the school), and 3 (4.3%) sessions missed when the participant was at school but unable to participate in the planned session (e.g., ill, dysregulated, improper footwear).

Two of the implementation strategies required adaptations that did not alter core components. First, the develop educational materials strategy was adapted during the trial to address challenges connecting two devices simultaneously. Troubleshooting identified a solution of connecting one device to a hotspot. Following FRAME-IS, this was a content modification involving the addition of elements to improve feasibility, informed by school staff implementing device use. Second, promote adaptability was expanded following the trial to allow flexibility in who could independently operate the device.

Schools structured device use as a physiotherapist intervention, requiring physiotherapists to operate or supervise use. This limited opportunities for device use and reduced therapy time for children not using the device. The adaptation was therefore a context modification with personnel implications, involving “loosening structure” so device use did not require physiotherapist oversight. The primary goal was to improve feasibility, with co-equal goals of increasing adoption and reducing inequities among students. This modification was informed by physiotherapists implementing device use.

#### Safety

Two minor adverse events were reported, which were both blisters on feet related to fit of ankle-foot orthoses that did not require any follow-up care or missed sessions.

### Effectiveness Outcomes

Each participant set three or four COPM goals, including one participation-based goal. Example COPM goals include: “Participate in upright activities in gym, playground, or outside 3 times per week” and “Use reciprocal stepping consistently for 30 seconds using regular walker”. Following the robot-assisting walking intervention, there was a mean improvement of 3.56 (SD 1.07, n=8) for COPM Performance scores and 2.84 (SD 1.17, n=8) for Satisfaction scores. Following the control intervention, mean improvement was 2.11 (SD 2.41, n=8) for Performance and 2.27 (SD 1.97, n=8) for Satisfaction. Changes in COPM scores are depicted in Figure 3.

**Figure 3.**
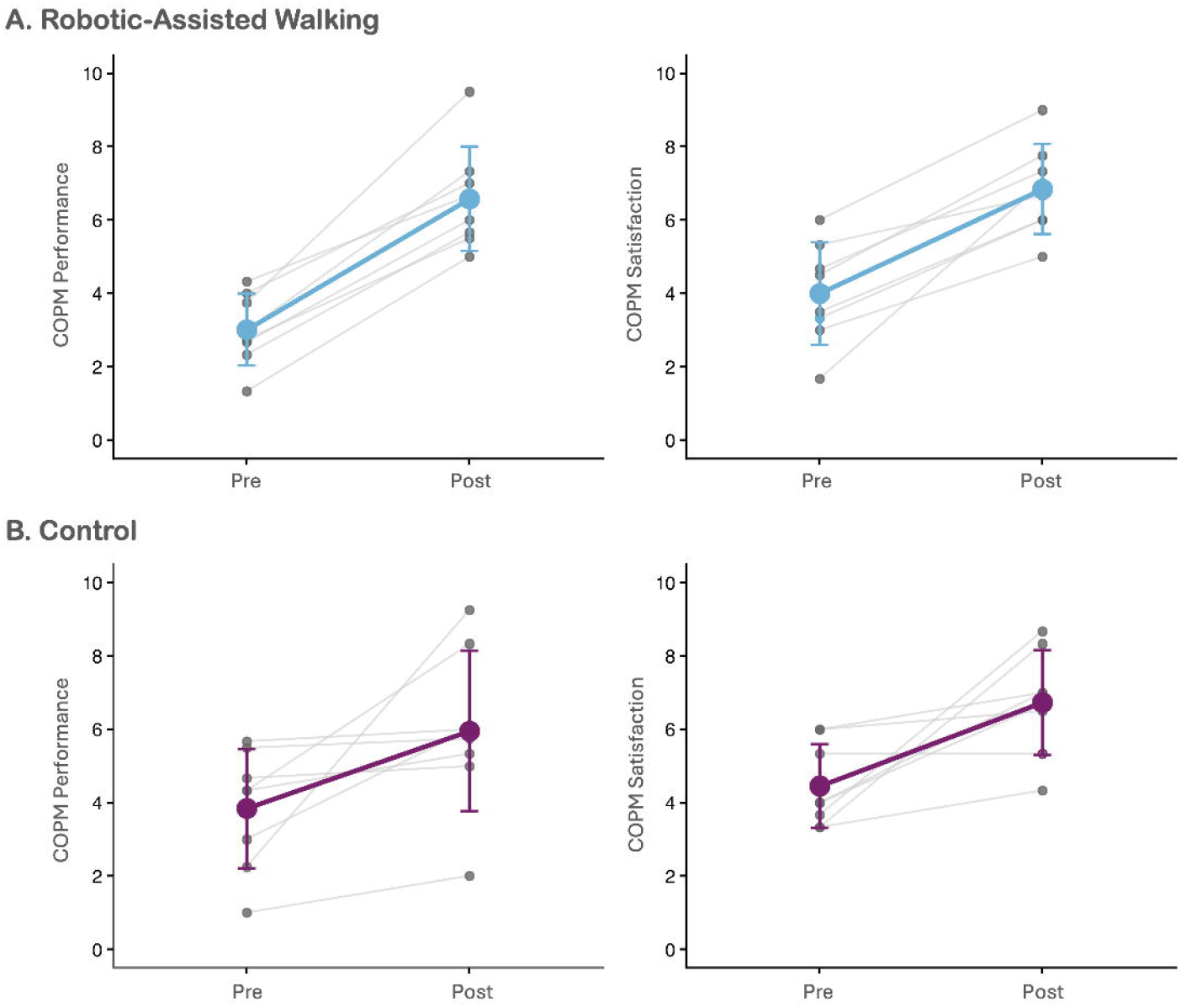
Individualized Participation-based Goals. **Caption**: Canadian Occupational Performance Measure (COPM) scores for (A) Robotic-Assisted Walking and (B) Control (usual device use). Grey markers and lines represent individual scores. Coloured markers and lines represent mean scores with SD error bars.

The COPM Performance treatment contrast was 1.45 (SD 1.97) points, corresponding to a moderate-to-large within-participant effect size of d_z_=0.74 (95% CI: -0.07 to 1.51). Power calculations indicated that 22 participants completing both periods would be required for the effectiveness aim of the definitive crossover design trial, corresponding to a minimum recruitment target of 28 participants after accounting for attrition and the randomization scheme. The between-condition correlation was r=0.59. A modest period effect of 0.59 points (period 1 minus period 2) was observed; therefore, period should be included a fixed effect in the definitive analyses, as is standard for crossover designs. No evidence of a sequence (carryover) effect was detected, although with four participants per sequence this pilot cannot meaningfully assess carryover. Finally, sensitivity analyses and Monte Carlo simulations supported a sample size of 22 completers.

Each child had 1 to 3 GAS goals per period. Mean GAS T-scores were 45.39 (SD 15.16, n=8) following the robot-assisted walking and 40.18 (SD 14.24, n=8) following control. A T-score ≥50 was achieved by one participant following both conditions, two additional participants following robot-assisted walking only, and one additional participant following control only. Example GAS goals include: “Participate in upright activities outside of the classroom 2x/week”, assessed using the YC-PEM, and “Increase walking distance in 2 minutes by 10 meters”, assessed using the 2MWT.

## Discussion

This pilot randomized crossover hybrid type 2 effectiveness-implementation trial provided preliminary evidence regarding both the potential effectiveness of robot-assisted walking and the viability of its implementation in schools for children with neurodevelopmental physical disabilities. These findings support progression to a definitive trial and provide evidence to optimize its design and conduct. This pilot provided encouraging evidence that use of robot-assisted walking devices may improve COPM Performance scores more than use of usual devices, although findings were inconclusive due to the sample size. The observed within-participant effect size was imprecise, as expected in a pilot, but together with the feasibility findings justifies proceeding to a definitive study. Both groups demonstrated a mean COPM score increase of ≥2 points, a threshold often used to indicate clinically meaningful change.^29^ However, this degree of improvement may reflect, in part, the ongoing developmental gains expected in this young population.^36^ Nevertheless, the pattern of greater improvement with robot-assisted walking warrants further investigation.

Modifications to implementation strategies focused on improving feasibility, which was the only implementation outcome that did not meet all pre-set criteria. Specifically, weekly target device use was not consistently met and therefore the team addressed technical and staffing issues that reduced device use. However, the most common reasons for reduced device use were participant absence or being unable to participate, which could not be mitigated. Research and school teams agreed that target use metrics were valuable for guiding implementation, rather than defining success.

Study design changes were identified for a future definitive trial. A 10-week intervention period is recommended and appears feasible, as all children in the pilot used the devices for at least nine weeks. Period 1 and 2 start dates should be planned with consideration for school holidays. Reducing the number of devices to one per school would address connection issues and reduce staffing demands. Although sharing one device among multiple children would increase access, sporadic use may not provide a sufficient dose to support meaningful improvements.^37^ Further, adjusting and charging devices between users was an identified barrier in our previous pilot study.^12^ Therefore, we recommend one device per school, with the device allocated to one child per intervention period to support consistent use and an adequate intervention dose.

Findings support retaining the COPM Performance score as the primary effectiveness outcome for the definitive trial, using the methods established in the pilot. Further, the success of data collection indicates that a decentralized trial is possible with measures collected at the child’s school or online. The measures selected for GAS goals should be expanded to include more participation-based options Although mean GAS scores improved from baseline, most participants did not achieve the expected level of improvement in either group. The predominance of motor function goals may have limited the ability of GAS to capture changes targeted by the participation-focused intervention. Furthermore, with novel interventions, determining the expected level of change can be difficult and our pre-defined thresholds for expected change may not have been achievable.

Additional secondary effectiveness measures should be included in the definitive trial to ensure that multidimensional outcomes are considered. The F-words for Child Development^38^ provide an approach to guide measure selection. While the F-words of Functioning and Fitness are assessed via the COPM and GAS, Friends and Fun could be assessed using an observational measure of social participation such as the Individual Child Engagement Record (ICER-R).^39^ Children’s perspectives could be captured in a simple study-built measure that supports multiple communication methods.

Implementation outcome measures should be expanded in a definitive trial to align with its broader implementation objectives. Given the complexity of implementing technology,^40^ understanding its value proposition and fit within school contexts will be important for successful implementation. Appropriateness, defined as the perceived fit or relevance of robot-assisted walking for meeting children’s participation needs within a school-based context,^41^ could be assessed alongside acceptability. The Intervention Appropriateness Measure (IAM) and Acceptability of Intervention Measure (AIM) would be easy to administer in school contexts.^41^ Acceptability could be further assessed using the Quebec User Evaluation of Satisfaction with Assistive Technology (QUEST).^42^ Collecting demographic data alongside an economic evaluation could be used to examine equity. Longer-term sustainability could be assessed using the Long Term Success Tool,^43^ while qualitative methods could provide a deeper understanding of school team and family perspectives.

Strengths of this pilot include the randomization, efficient type 2 hybrid design, real-world setting, and prospective collection of implementation data. Limitations include the small sample, which reduces certainty but was a responsible use of resources for a pilot trial. There were less sites than will be included in the definitive trial and all sites had previous experience with robot-assisted walking devices, which may have biased results. Our engagement of family partners and school physiotherapists undoubtedly supported successful implementation. However, we did not evaluate engagement, which could have been done using the Public and Patient Engagement Evaluation Tool (PPEET).^44^

Overall, this pilot trial demonstrated the potential for robot-assisted walking to be implemented in school settings while providing preliminary evidence of benefits for student participation. Findings regarding intervention delivery and implementation will inform refinement of the design of a future definitive trial.

## Data Availability

The datasets used during the current study are available from the corresponding author on reasonable request with evidence of ethical approval.

## Acknowledgements

We thank the children and families for dedicating time and effort to participate in this study. We also thank the school teams for their collaboration and contributions throughout the study.

