## Supplemental Material for "Upright Participation for Learners with Physical Disabilities: A Pilot Randomized Crossover Effectiveness-Implementation Trial of Robot-assisted Walking in Schools"

This supplement has been provided by the authors to give readers additional information about the work.

### Table of Contents

***Additional Details: Methods for Goal Attainment Scaling ...................................................... 2***

**Additional Details: Methods for Goal Attainment Scaling**

Goal Attainment Scaling (GAS) was based on the scoring characteristics of each measure. For example, the Young Children's Participation & Environment Measure (YC-PEM) was used to assess participation frequency.[1] The items (A1, A2 and/or A3) that reflected participation in an upright position were selected based on the child's COPM goal.[1] The school staff member most involved with the participant scored their participation frequency on this item using the YC-PEM's 8-point scale: from never (score 0) to once or more each day (score 7) via an emailed questionnaire. GAS level 0 represented the target change while each GAS level represented a 1-point difference on the YC-PEM scale in participation frequency (see Figure x for an example of this scaling).

The Early Clinical Assessment of Balance (ECAB)[2] was used to assess static and dynamic postural control across 13 items (maximum of 100 points) and was videotaped for subsequent scoring by a trained research clinician (CD). ECAB use for GAS was tailored to the child and COPM goal. For example, when use of the entire ECAB and its total ECAB score was appropriate to the goal, each GAS level set represented a 5-percentile change based on age- and Gross Motor Function Classification System (GMFCS)[3]-matched norms. When individual ECAB items were more appropriate, a 1-point change in the relevant sub-score represented one GAS level.

The two-minute walk test (2MWT)[4,5] was used to assess walking endurance based on the distance walked in 2-minutes using their usual gait aid, as recorded by the school team. For the 2-MWT, change from level -2 to 0 (expected level of change) represented what was felt to be the minimal clinically important distance difference for that child.

When these measures did not align with the COPM goal, individualized tasks were chosen for the GAS goal. Possible tasks were timed standing, timed sitting, and the number of steps initiated during 5-minutes when using a manual walker. For these individualized tasks, GAS levels were established based on the clinical judgment of the school physiotherapist.
